# Loneliness and the onset of new mental health problems in the general population: a systematic review

**DOI:** 10.1101/2021.01.26.21250587

**Authors:** Farhana Mann, Jingyi Wang, Ellie Pearce, Ruimin Ma, Merle Schleif, Brynmor Lloyd-Evans, Sonia Johnson

## Abstract

**Background:** Loneliness is associated with poor health including premature mortality. There are cross-sectional associations with depression, anxiety, psychosis and other mental health outcomes. However, the direction of causation is unclear and clarifying the evidence from longitudinal studies is a key step in understanding this relationship.

**Aims:** We synthesized evidence from longitudinal studies investigating the relationship between loneliness and new onset of mental health problems, in the general population.

**Method:** We systematically searched six electronic databases, unpublished sources and hand-searching of references, up to March 2020. We conducted a meta-analysis of eight independent cohorts, and narrative synthesis of the remaining studies.

**Results:** We included 20 studies, of which the majority focused on depression. Our narrative synthesis concluded that loneliness at baseline is associated with subsequent new onset of depression. The few studies on anxiety also showed an association. Our meta-analysis found a pooled adjusted odds ratio of 2.33 (95% C.I. 1.62 – 3.34) for risk of new onset depression in adults who were often lonely compared with people who were not often lonely. This should be interpreted with caution given evidence of heterogeneity. Most of the studies were in older adults.

**Conclusion:** Loneliness is a public mental health issue. There is growing evidence it is associated with the onset of depression and other common mental health problems. Future studies should explore its impact across the age range, look beyond depression, and explore the mechanisms involved.

## Introduction

Mental illness remains a leading cause of disability worldwide, and the COVID19 pandemic may mean a further increase in its burden across the population (1, 2). Social relationships and loneliness are important candidate areas for preventive interventions. Loneliness can be defined as a distressing mismatch between the quantity and/or quality of social relationships a person has, and what they desire(3). It is related to concepts such as social capital, objective social support and social isolation (4), but is conceptually distinct. Loneliness relates specifically to the perceived quality or quantity of social relationships, as opposed to objective assessments of them. Population surveys have suggested high levels of loneliness amongst both older and young adults (under 25), giving a roughly ‘U-shaped’ distribution (5, 6). Those with prolonged mental health problems and/or receiving psychiatric treatment are at greatest risk of ‘severe’ loneliness (highest scores on loneliness scales). With the possibility that causality between loneliness and mental health problems could be in either direction or both, synthesizing the longitudinal evidence is an important way to clarify this.

Cross-sectional associations have recurrently been found between loneliness and several mental health problems, including depression (7), anxiety (8), personality disorder (9), psychosis (10) and suicidal ideation (11). An important consideration has been the crossover between loneliness and depression as concepts. A number of studies have demonstrated these as partially correlated but distinct, and they may in fact share a reciprocal relationship (12).

A 2016 systematic review concluded that there is an association between poor social support and depression but did not search for studies on loneliness, or conditions other than depression (13). This is a significant gap as the specific subjective experience of loneliness has been demonstrated as having an important independent effect on health (14). Of note, the review found that emotional support in particular (conceptually closest to loneliness) was most commonly associated with protection from depression. A more recent meta-analysis on loneliness did not include a systematic literature search, scanned the literature only for the terms depression and loneliness, and crucially did not look for longitudinal studies (15). To address these gaps, we ask: ‘does loneliness lead to the onset of mental health problems in the general population?’

## Method

### Search strategy and selection criteria

This review reports on studies of adults in the general population (i.e. non-clinical cohorts), that address the question of whether baseline loneliness is associated with the later onset of mental health problems. We included a wide range of mental health outcomes including depression, anxiety, bipolar disorder, personality disorder, and psychosis (full list of terms in supplementary material 1). In this report, our exposure of interest was ‘loneliness’. However, given that there are a number of related subjective social relationship concepts, we set out to include a broader range of terms in our original search. This was to ensure we did not miss studies that looked at both perceived social support and loneliness as main exposures independently, but may only have listed ‘social support’ in their title or key words.

We did not include studies investigating people with intellectual disabilities, children under the age of 16, people with organic mental illness, cohorts of people selected on the basis of a primary physical health diagnosis, or where loneliness and mental health problems were not the primary exposures and outcomes respectively. The review was registered on PROSPERO under registration number CRD42015014784.

We initially systematically searched six electronic databases up to and including June 2018. We later updated the searches, using the exact same key words and databases to include studies up to March 2020. The databases searched were: Medline, PsychINFO, EMBASE, Web of Science, CINAHL and the Cochrane Library. No language or publication period restrictions were applied. The reference lists of included studies were hand-searched, as well as references listed in relevant review papers. We also searched for dissertations, conference reports or other non-published reports, on both Zetoc and OpenGrey databases. Where relevant, we contacted study authors for further data (if it was likely to mean further papers could be included in a meta-analysis for example). We received responses from three authors in this regard.

Searches were conducted using both subject headings (MeSH terms) and text words within titles and abstracts. Search terms were adapted as required for different databases. The initial search was conducted as part of a broader search to include longitudinal studies on loneliness in people with established mental health problems (for a separate, related review (16)). Our searches combined terms for ‘loneliness and related concepts’, ‘mental disorder’, and ‘onset’. A fuller list of search terms is provided in the supplementary material 1.

All identified titles were screened electronically by independent reviewers (FM and JW for the initial search, and EP, MS and FM for the update). The abstracts of relevant papers were read, and full texts retrieved if the abstract suggested they were likely to meet our inclusion criteria. All full texts of studies included by one assessor were checked by a second, to ensure they matched our criteria. Any queries relating to inclusion/exclusion were resolved through discussion with a third reviewer (SJ).

### Quality Assessment

We used the Mixed Methods Appraisal Tool (MMAT) Version 2011 to structure our quality assessments (17). This is a tool specifically designed for methodological quality appraisal across a range of study types (both quantitative and qualitative). The tool provides specific criteria against which to assess quality for each type of study. For our purposes, we used the quantitative, non-randomised domain designed for cohort studies. Papers are rated across the following four domains: selection bias, measurement quality, adjustment for confounders, and percentage of complete outcome data/response rate/follow-up rate. Scoring the papers on each category gives an overall rating ranging from ‘*’ to signify poorest methodological quality (one criterion met) to ‘****’ (all four criteria met).

Given the focus on the general population, we also included additional quality items from the Newcastle-Ottawa Scale (NOS) (18). This is a tool developed to assist with the evaluation of non-randomised studies, and we included two questions that ask about representativeness.

A sample of quality assessments and data extraction findings (20%) were rated independently by independent reviewers (FM, JW and MS) to assess consistency.

### Narrative and statistical analysis

We aimed to consider all included studies in a quantitative meta-analysis, but were aware it might not be possible to combine them all due to variation in statistical approach, and reported results. To provide a meaningful quantitative synthesis, we pooled results from studies that provided adjusted quantifiable estimates for the risk of mental health problems. The statistical analysis was carried out using STATA v15·1, and the pooled odds ratios using random effects are reported. We measured heterogeneity between studies using the chi-squared test and the I^2^ statistic. We needed at least three independent cohorts to be eligible for meta-analysis to carry it out.

For the remaining studies, we provide a narrative synthesis, guided by the principles in Popay et al (2006) (19). We considered how any relationship between loneliness and mental health differed by important characteristics such as age and gender, as well as quality.

## Results

The results from our search are represented in the Preferred Reporting System for Systematic Review and Meta-Analysis (PRISMA) flow diagram, which presents the total number of studies up to March 2020 (Figure 1). Our original search retrieved 15154 non-duplicate records. After screening titles and abstracts 879 full text articles were assessed for eligibility. 800 studies excluded by one reviewer were screened independently by a second, and agreement was over 99%. Sixteen studies on loneliness and the onset of mental health problems, or on a combination of onset and outcomes in the general population, were included. Four further studies were identified through reference chaining, giving a total of 20 studies.

**Figure 1.**
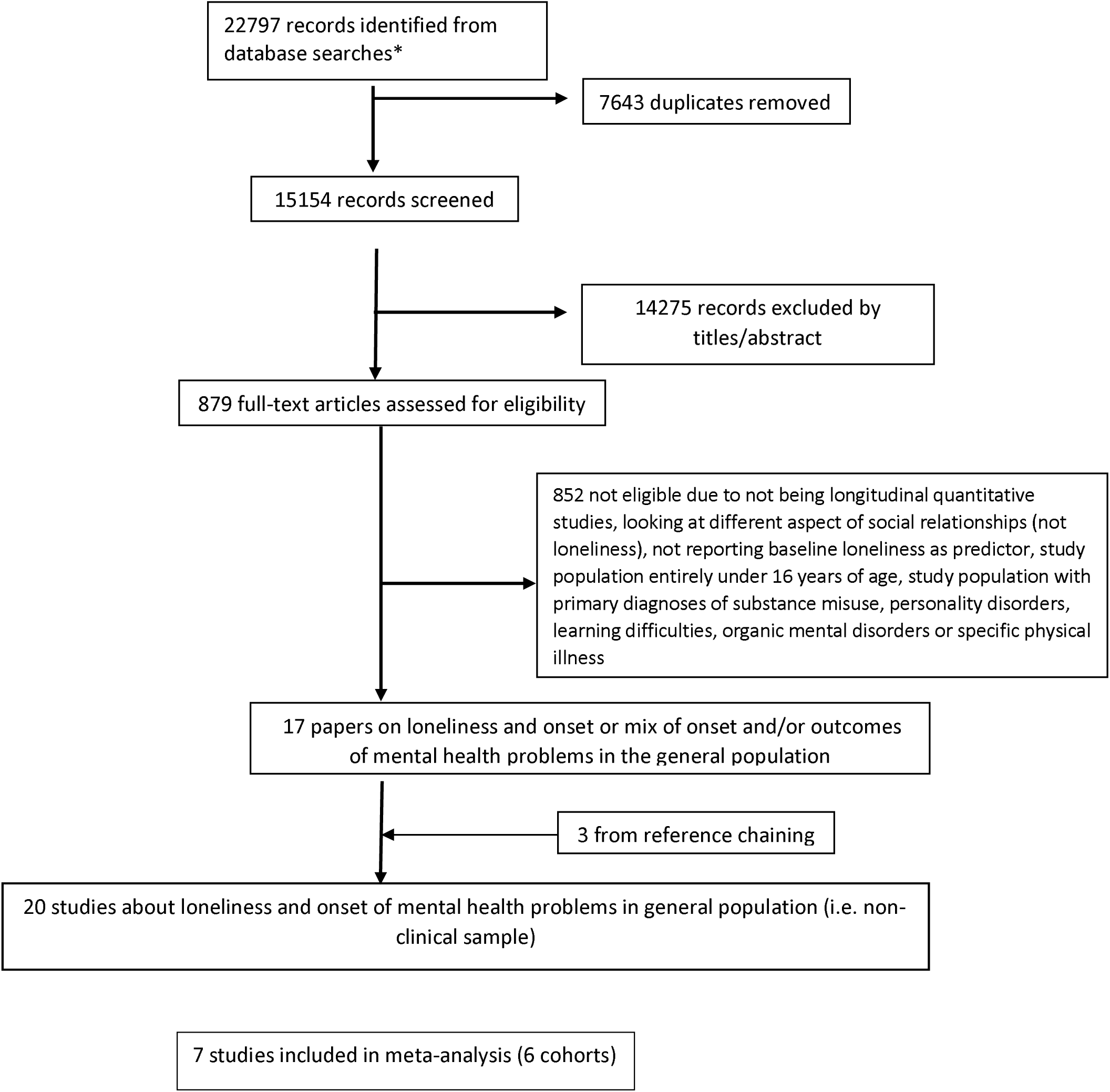
PRISMA flow chart to show search strategy. * search terms covered loneliness and perceived social support, and both outcome and onset of mental health problems. These numbers include updated searches up to and including March 2020.

The main characteristics and results of all 20 studies are summarized in Table 1. Overall, nearly all of the results suggested baseline loneliness to be associated with the onset of depression or anxiety. There was considerable variation in size, ranging from 83 to 22,268. In total, accounting for studies reporting data on the same cohorts, the included papers covered 84, 188 people across ten countries. The length of follow-up ranged from 6 months to 16 years, with a mean of 3.5 years. A large majority of studies (nineteen out of 20) included the onset of depression or depressive symptoms as a primary outcome, with one looking at both depression and anxiety (20), and another exclusively at anxiety disorders (21). One study reported on ‘common mental health problems’ (items that included a combination of mood and anxiety symptoms from the ‘CORE-GP’ questionnaire). The majority (fourteen) focused on people aged over 50 (eight studies in people over 65), with the remainder covering new mothers or younger adults including university students. Nearly all of the studies were from the US or Europe.

**Table 1.**
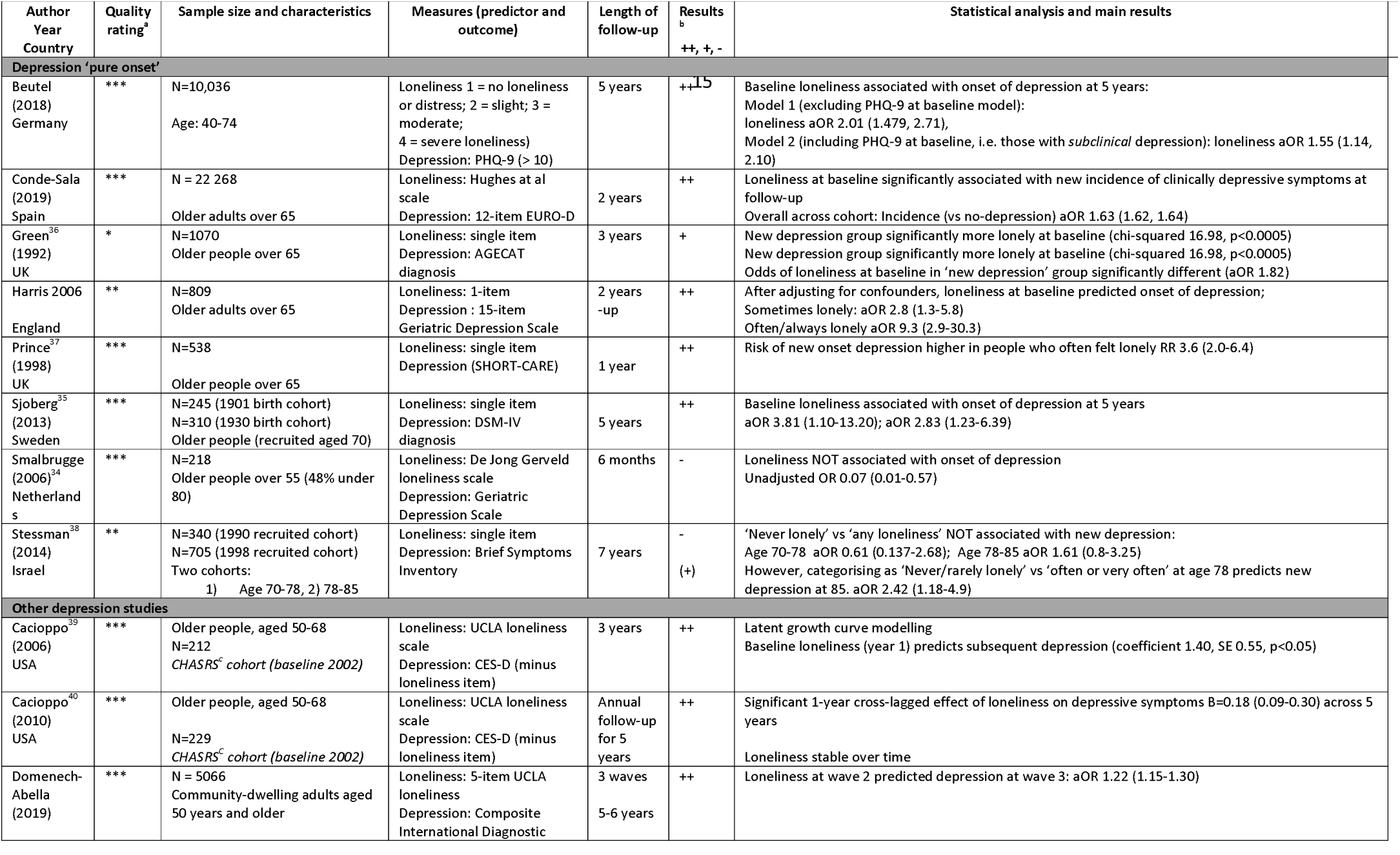
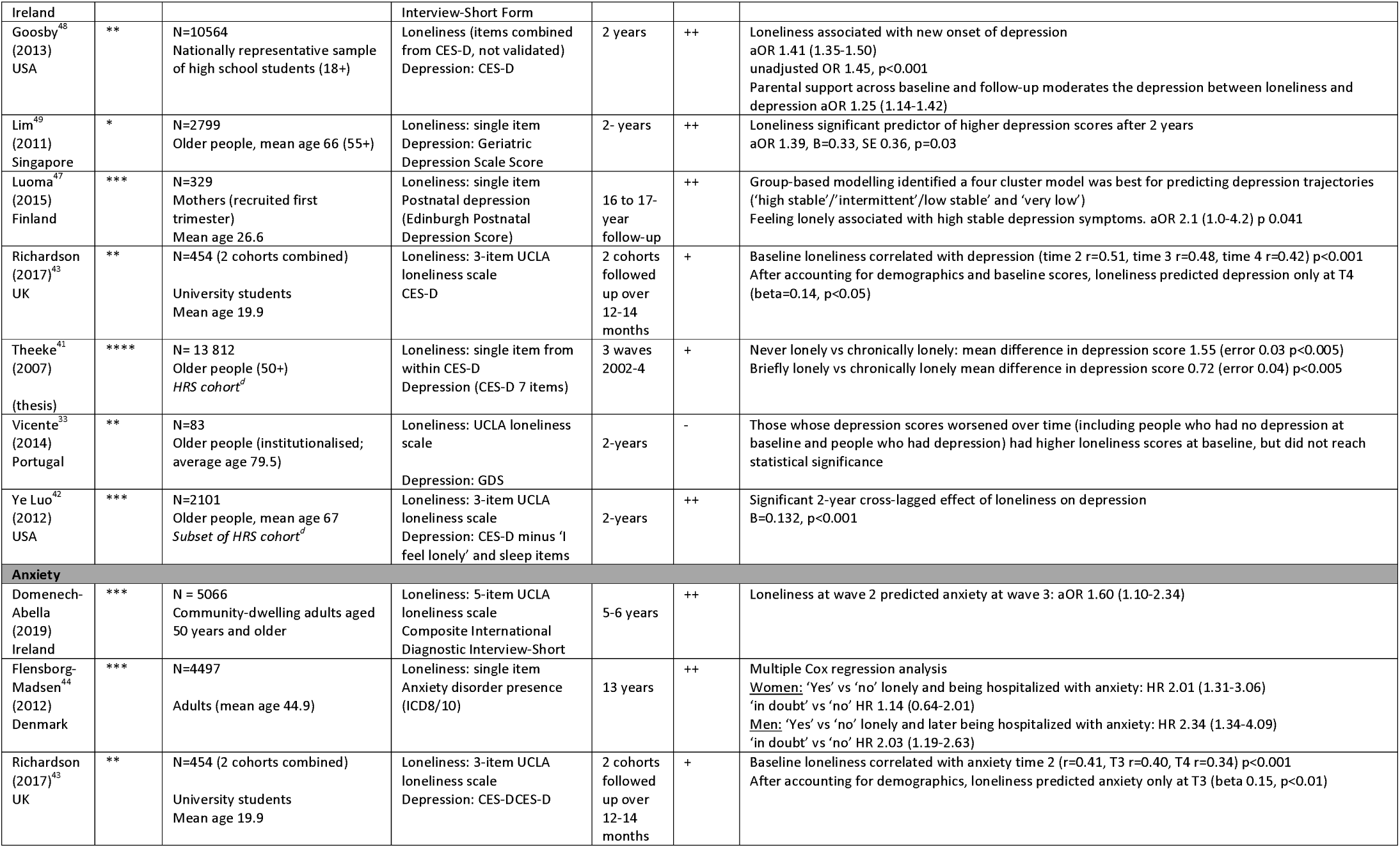
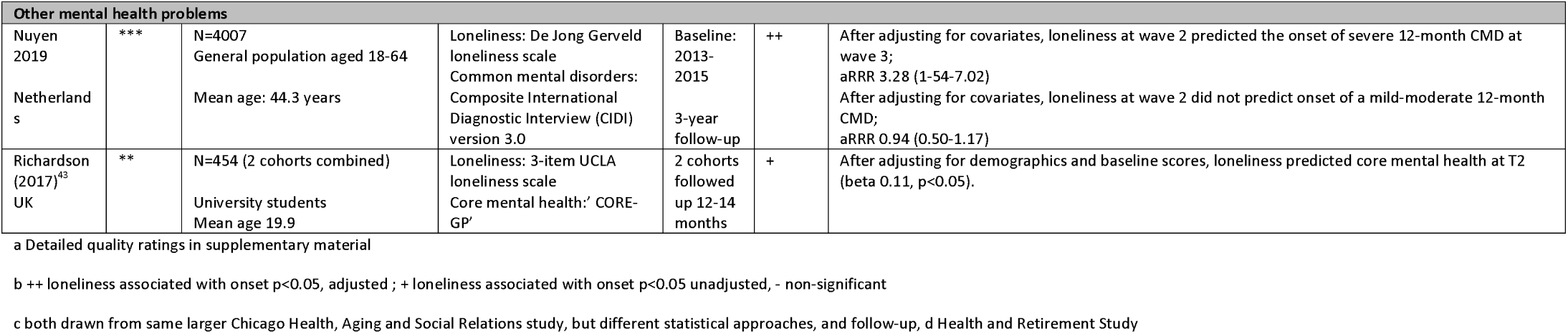
Summary of main findings from included studies across depression, anxiety and ‘core mental health’.

Only one study required translation, from Portuguese to English (22). Eight studies (all specifically on depression) took steps to screen for, and remove, all people who already had depression at baseline. We referred to these as ‘pure onset’ studies (23-29).

Two papers analysed data from the Chicago Health Ageing and Retirement Study (CHASRS) cohort, but were independent studies that used different statistical approaches and reported on different follow-up periods (30, 31). We have included both sets of findings in the narrative synthesis but neither gave results that could be combined in a meta-analysis. In addition, a PhD thesis investigated loneliness in people from the Health and Retirement Study (HRS) (32) cohort, while a different study also reported on the same cohort independently (33). Again, both sets of results are described, but neither contributed to the quantitative analysis due to differences in methodology and statistical output.

The most commonly used validated loneliness measure was the UCLA Loneliness scale (both 20-and 3-item versions), used in seven studies(34). Three studies used the De Jong Gierveld Scale (scoring >3 was classified as ‘highly’ lonely), and the remainder used composites of relevant items in other tools (e.g. CES-D) or single items on loneliness (details in Table 1). There was variation in how loneliness was categorised (e.g. dichotomised versus continuous score).) The quality ratings of included studies ranged from the lowest ‘*’ to excellent ‘****’, with the majority being rated ‘moderate to good’. With regard to representativeness, a number of studies took steps to use e.g. multistage probability designs to boost inclusion of ethnic minority groups, or national register data but others were unclear on the steps in the selection process. Quality was also affected by loss to follow-up. Rates of depression onset were low in some studies, which may mean an underestimation of the true effects on this outcome. Details on individual quality ratings are given in the supplementary material 2.

## Meta-analysis

We performed a meta-analysis of all studies that provided adjusted odds ratios for loneliness and risk of depression. This gave a total of seven eligible studies. One study reported on two independent cohorts (1901 vs 1930 births)(24), resulting in inclusion a total of eight distinct cohorts. A random-effects analysis was chosen given that the individual studies sampled people from distinct populations. In order for the combined result to be meaningful, we combined studies that used a binary loneliness measure, but also provide a result for three studies that use continuous loneliness measures in supplementary material 3.

The result across all eight independent cohorts was that the (adjusted) odds of depression was higher in people who were often lonely compared with those who were sometimes or never lonely. The pooled odds ratio for loneliness being associated with subsequent onset of depression was 2.33 (1.62 – 3.34). This should be interpreted with a degree of caution given the I^2^ statistic was moderately high at 64%.

**Figure 2.**
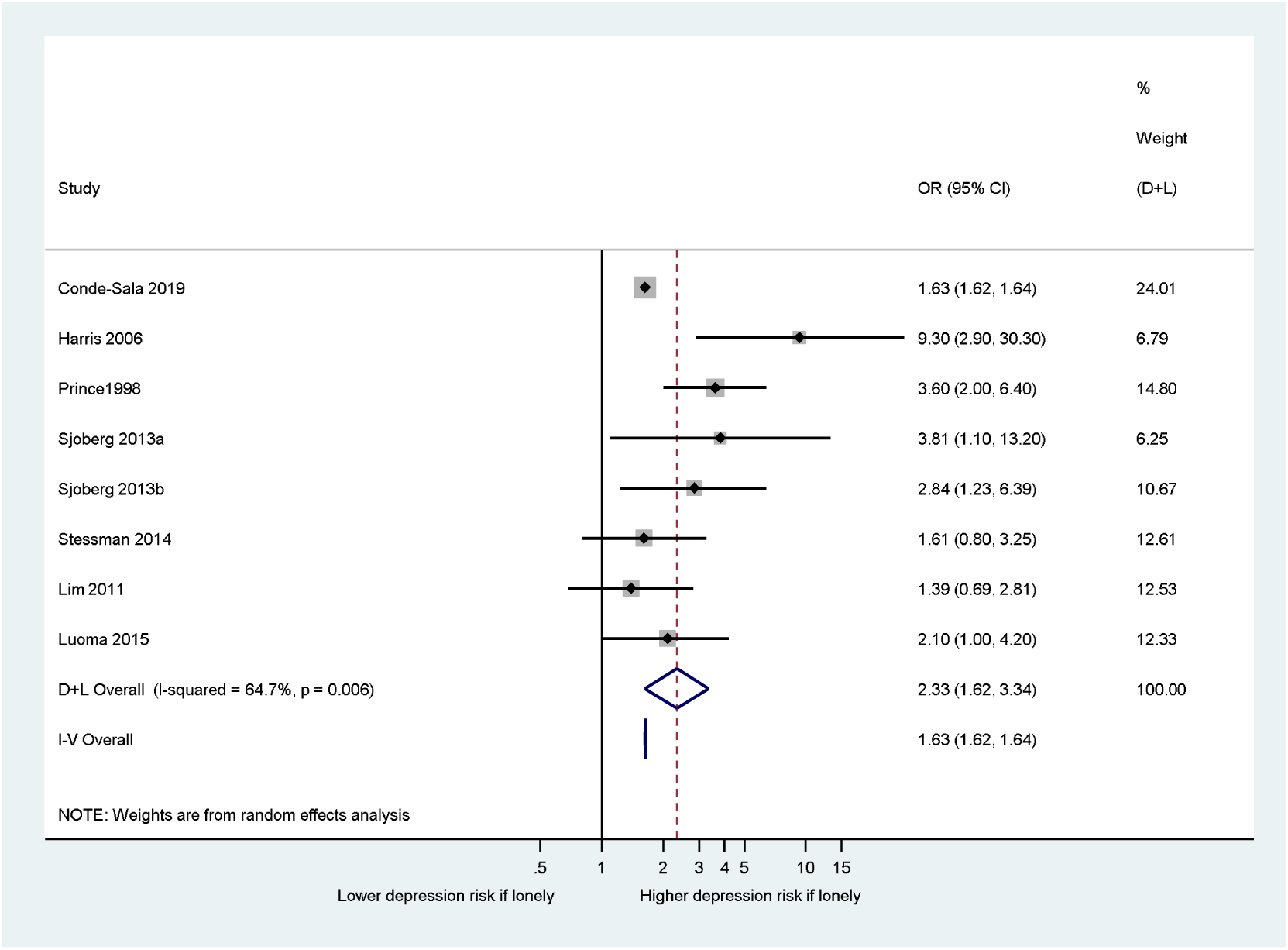
Forest plot to show association between loneliness and new onset of depression (loneliness as binary independent variable).

Five of the seven studies meta-analysed with binary loneliness outcomes, were ‘pure’ onset studies. That is, all participants with current or past depression at baseline were excluded at the outset so any depression at follow-up was entirely new. Six of the studies were in older adults, with the remaining one looking at new mothers. All used a range of categorical classifications of loneliness but not validated tools. The newest study (29) was the largest (n=22 268), follow-ups ranged from 6 months to 17 years, and most of the studies were of good quality (five rated ‘***’ in our quality rating). The majority of studies adjusted for gender. One study took steps to adjust for the more objective baseline measure of ‘number of contacts’(24), while others also made adjustments for objective markers such as living alone and other domains of social support (25, 26).

### Narrative synthesis

Two remaining ‘pure onset’ studies both favoured an association between loneliness and depression (RR 3.6 (35) and aOR 1.82 (36), but did not provide results that could be meaningfully combined in a meta-analysis. In one case (36), the lack of confidence interval precluded inclusion in the meta-analysis. The author was unable to provide that information as the original data was no longer available.

The characteristics and main results of these studies are summarised in Table 1. The results nearly all showed a significant association between baseline loneliness, and subsequent onset of depression (or anxiety).Two studies (33) (32) analysed data from the HRS -a national longitudinal panel study of health and ageing in the US (in people aged over 50) with a stratified multistage probability design. Both suggested an association between loneliness and depression, but each used different combinations of items to measure loneliness and different statistical approaches. One of the studies reported coefficients from a 2-year cross-lagged panel analysis, and the other provided mean differences in depression score.

Both Cacioppo et al (2006)(30) and Cacioppo et al (2010)(31) analysed data from the CHASRS – a nationally representative US longitudinal study on health and social relationships in people aged 50-68. Both studies had a baseline year of 2002, but differing lengths of follow-up and statistical approaches. The 2006 study used latent growth curve modelling, and suggested baseline loneliness predicted depressive symptoms at three-year follow-up (coefficient 1·40, SE 0·55, p<0·05), while the second study reported a significant one-year cross-lagged effect of loneliness on depression across five years (B=0·18 (0·09-0·30)). This effect remained consistent when accounting for social networks, neuroticism and demographic factors. The 2006 study found depression also predicted a steeper rise in loneliness levels over time, suggesting a reciprocal relationship.

Vicente et al (37) was the smallest study, and the only one to not show any significant association between loneliness and depression. It did suggest people whose loneliness scores worsened over time also had worsening depression scores (compared with people with stable or improving depression scores) but this did not reach statistical significance. Finally, Richardson et al(20) studied loneliness in 454 students (mostly white females). Baseline loneliness score was correlated with depression at all four follow-up time points. However, once baseline depression scores and demographics were adjusted for, loneliness only predicted depression at the final follow-up (twelve to fourteen months, beta=0·14, p<0·05).

## Studies on loneliness and anxiety

Three studies included measures of anxiety as key outcomes, with loneliness as the exposure. Flensborg-Madsen et al (21) followed a cohort of Danish adults (mean age 45) over thirteen years. Compared with answering ‘no’ to being lonely at baseline, women who responded ‘yes’ had HR 2·01 (1·31-3·05), and those who responded ‘in doubt’ had HR 1·14 (0·64-2·01). In men, the corresponding HRs were 2·34 (1·34-4·09) and 2.03 (1·19-2·63). People with anxiety at baseline were screened out, making this a ‘pure onset’ study. Domenech-Abella (2019) (38) found loneliness in one wave was associated with onset of anxiety in the next wave (aOR 1.60, CI 1.10-2.34).

Richardson et al(20) also studied anxiety and ‘core mental health’ as outcomes. Baseline loneliness correlated with anxiety at three months (r=0·41) six months (r=0·40), and twelve months (r=0·34,) p<0·001. However, once adjusted for baseline demographic variables, loneliness was only associated with anxiety at six months (beta 0·15, p<0·01).

## Loneliness and other mental health problems

In the same study (Richardson et al) (39), loneliness was associated with ‘core mental health’ (mix of mood and anxiety items defined by the CORE-GP questionnaire) at follow-up time 2 (approximately six months), but not at other follow-up time points.

A Dutch study (40) reported ‘common mental disorders (CMD’), classified as mild, moderate and severe. CMD included mood disorders (including bipolar I) and substance misuse. It showed loneliness was significantly associated with onset of ‘severe’ CMD after 12 months (aRR 3.28, 1.54-7.02), but not with mild-moderate CMD.

We found no longitudinal studies with onset of psychosis or personality disorder as outcomes.

## Discussion

This is the first systematic review of quantitative longitudinal studies, addressing whether loneliness is associated with the subsequent onset of mental health problems in the general (non-clinical) population. There is growing international interest in the health impacts of feeling lonely. This review focused specifically on the subjective feeling of loneliness, as opposed to a broader range of related but distinct concepts often grouped together under the umbrella of ‘social relationships’.

We established that the odds of developing new depression in adults, is more than double (pooled adjusted OR 2.33) in people who are often lonely compared with those who are not/rarely lonely. This is after adjustment in individual studies for various factors – including sociodemographic variables, functional limitations, and physical illness. Several of the studies took steps to adjust for other social relationship measures such as social support and found the effects of loneliness persisted. This is to be interpreted with a degree of caution given the heterogeneity of findings, but all included studies showed a positive association. A smaller study suggested it is possible that occasional experiences of loneliness may not in themselves lead to depressive symptoms, but more frequent or persistent loneliness might be more of a concern (41). Understanding at which point loneliness becomes a stronger predictor of depression will be important in developing interventions.

The remaining studies on depression (outside meta-analysis) also nearly all favoured loneliness being associated with an increased risk of depression onset over time. A finding of note was that two studies suggested a reciprocal relationship between depression and loneliness (42, 43). Whilst many of the studies used single item or other categorical classifications of loneliness, there was broadly no notable difference in findings between these and studies using validated loneliness measures. The choice of depression measure did not show any pattern with regard to results either. Most studies were of ‘moderate’ to ‘good’ quality (2/3 stars out of 4) and the largest and best quality studies showed results consistent with our main conclusion that people who are lonely are at greater risk of becoming depressed.

We found a general lack of longitudinal studies demonstrating the effects of loneliness in younger adults. New mothers are a group vulnerable to loneliness as well as mental health problems, and lack of social support is broadly known to be associated with postnatal depression (44). In the one study included in this population, antenatal loneliness was associated with people scoring high for depression up to seventeen years later (45). Two further studies that focused on younger adults, also concluded loneliness was associated with depression up to eight years later (20, 46).

A large study with thirteen-year follow-up (47) suggested loneliness is associated with the onset of anxiety. There was also evidence loneliness predicted the onset of common mental health problems in adults.

Of note, there were no studies of onset of psychosis or personality disorder identified in the search. A recent review of loneliness in people with existing psychosis noted a lack of rigorous studies(10). Existing work in this group of people is limited to predominantly cross-sectional studies (48), partly down to the relatively lower incidence of psychotic illness.

With regards to gender, most studies that adjusted for it, or looked for interaction, did not observe any significant effects. The wider literature has not suggested a consistent picture with regard to differential effects by gender. Also of note, several of the studies took steps to adjust for other social relationship measures such as social support, and found the effects of loneliness persisted. Most studies did not provide subgroup analysis by ethnic origin, but some of the cohorts studied included study populations actively recruited to represent e.g. Hispanic or Black minority groups in the US. There were no clear patterns identified in terms of ethnicity, though cross-sectional work has highlighted ethnic minority status as a risk factor for being lonely in the general population (6).

## Future Research

While the overall message is that loneliness appears to be associated with onset of depressive and anxiety symptoms in the general population, this review has highlighted several important research gaps. Firstly, few studies included main outcomes other than depression. There was no work on onset of other mental health problems such as psychosis or personality disorder, despite cross-sectional evidence loneliness is a concern in these groups (49) (8).

There was also a notable imbalance in the number of studies on older people compared with other age groups. Given there is a peak in loneliness in young adulthood (5), which also happens to be a peak time for the onset of several mental health problems, this is an important gap. There are likely to be differences in what drives loneliness, and/or how it impacts on health in people aged under-25 compared with those in their 70s or 80s (50). This information will also be important in developing appropriate, effective interventions to reduce loneliness across the age range.

Any association between exposure and outcome raises questions around mechanisms. There is a body of research into a number of different potential mechanisms through which loneliness may lead to poorer health, predominantly poorer physical health, including cardiovascular mortality. There is evidence for altered immune system function (51), changes in hypothalamic-pituitary axis function and differences in cortisol level (52), other physiological changes such as raised total peripheral resistance, as well as poor sleep and altered health behaviours. It is possible some of these may be on the causal pathway (if indeed a causal association exists) between loneliness and e.g. depression. The finding that depression in turn increases feelings of loneliness suggests a complex interplay between these experiences. Future studies of loneliness and mental health will benefit from factoring in measures of depressive symptoms, and depression has been suggested as a mediator between loneliness and poor physical health.

Cognitive mechanisms involved may also be different depending on what has led to the loneliness – physical disability versus social anxiety or bereavement, for example. There are a number of potential cognitive, behavioural and social factors that may be more relevant in people who are lonely versus those who are not. University students who identify as lonely tend to make more negative appraisals of how other people perceive them in actual and anticipated social circumstances (53), leading to social withdrawal and reduced informal social support. People who are more lonely also tend to engage in more ‘unhealthy’ lifestyle behaviours. It has been hypothesized internalised stigma or excessive awareness of threat in social situations may be relevant in developing psychosis, and lonely people tend to experience these thought processes more than those who are not lonely. The experiences of people with learning disabilities or autism may also require more focused study, amongst other groups. Future research could also explore when and how loneliness becomes persistent and severe enough to cause mental health problems. Such knowledge will be important for potential strategies at alleviating loneliness.

There is prior evidence autonomy and a sense of perceived control over one’s circumstances (in terms of loneliness/isolation) can be protective for people experiencing loneliness (54), and early studies on social support have suggested the effects on mental health can vary.

## Policy Implications

Future policy in this area will need to consider the experiences of people across the age range, and recognise the impact loneliness has on future mental health. We have previously discussed the different levels at which interventions to target loneliness could be aimed, i.e. individual, local community, and wider society (55). We have also discussed the importance of considering both direct and indirect interventions to address loneliness in the general population (the latter including transport, housing, and tackling poverty for example). The findings in this review further underline the importance of prioritising loneliness across these policy areas, and sit alongside existing evidence that it is associated with poor physical health outcomes.

Studies on loneliness interventions in people with established mental health problems have tended to demonstrate only modest effect sizes when they show any impact on loneliness (56), though the number of studies is small. One consideration in light of this, along with the findings from our review, is that there is also benefit intervening proactively through public mental health initiatives. Raising the profile of loneliness as a legitimate health concern is a start, and the future of interventions may include approaches including social prescribing (e.g. taking referrals from primary care) and community-level interventions(55). Public Health England previously published estimated cost savings if loneliness in older people were to be reduced. This is likely a gross underestimate of the cost savings if loneliness were to be tackled across the age range. De Jong Gierveld has discussed the potentially superior effects of interventions that highlight a person’s need to invest in their ‘social convoys’ throughout life, before they may find themselves both unwell and lonely (57).

In the context of the COVID19 pandemic, these findings highlight the need to proactively promote community connections and prevent loneliness as they are likely to yield significant health benefits down the line.

## Limitations

Despite its strengths, there are a number of limitations to this review. Firstly, the focus was on adults of all ages – but including children would give an even broader understanding of the life course perspective, and earlier effects on mental health. Whilst our aim here was to be specific to loneliness, the inter-relationships between the different ‘social relationship’ concepts remains an important area of research, though beyond the scope of this particular review. In the quantitative analysis, we took steps to include studies looking at similar populations, investigating loneliness and depression. However, there were some differences in terms of the specific covariates adjusted for (Table 1), which must be kept in mind when interpreting the pooled odds ratio.

The quality of included papers is of course an important consideration, and most papers included were of moderate quality. Less than half used well-established validated loneliness measures. We recognise the vast majority of studies included here originate in higher income countries, and there is some evidence cultural context (e.g. individualism versus more collectivist settings) could be an important consideration in how loneliness may impact mental health (58). Rates of depression onset were low in some studies, which may mean an underestimation of the true effects on this outcome.

## Conclusion

This review indicates adults who experience more loneliness in the general population, are at more than twice the risk of developing depression over time. There is more evidence in older adults, but longitudinal studies suggest increased risk of depressive symptoms in younger age groups also, including new mothers and university students. There is also evidence loneliness is associated with increased risk of anxiety disorder, but a lack of research exploring the effects on onset of other mental health problems like psychosis. A number of important research gaps and future priorities have been identified. We will benefit from a better understanding of mechanisms to support the development of effective interventions. This review suggests loneliness is a significant public mental health concern.

## Supporting information

Supplementary Material 1 Search Terms

Supplementary Material 2 Characteristics Table Detail

Supplementary Material 3 Quality Ratings

Supplementary Material 4 Forest Plot Continuous

Supplementary Material 5 Depression and Loneliness Rates

## Data Availability

Data from the review can be requested (STATA files of meta analysis, original searches and EndNote for references)

## References

1. Pierce M, Hope H, Ford T, Hatch S, Hotopf M, John A, et al. Mental health before and during the COVID-19 pandemic: a longitudinal probability sample survey of the UK population. Lancet Psychiatry. 2020; 7(10): 883–92.

2. Mehta N, Davies SC. The importance of psychiatry in public mental health. Br J Psychiatry. 2015; 207(3): 187–8.

3. Peplau L. Perspective on loneliness. In: Loneliness: a sourcebook of current theory, research and therapy John Wiley and Sons, 1982.

4. Wang JY, Lloyd-Evans B, Giacco D, Forsyth R, Nebo C, Mann F, et al. Social isolation in mental health: a conceptual and methodological review. Social Psychiatry and Psychiatric Epidemiology. 2017; 52(12): 1451–61.

5. Victor CR, Yang KM. The Prevalence of Loneliness Among Adults: A Case Study of the United Kingdom. Journal of Psychology. 2012; 146(1-2): 85–104.

6. Lasgaard M, Friis K, Shevlin M. “Where are all the lonely people?” A population-based study of high-risk groups across the life span. Social Psychiatry and Psychiatric Epidemiology. 2016; 51(10): 1373–84.

7. Barger SD, Messerli-Burgy N, Barth J. Social relationship correlates of major depressive disorder and depressive symptoms in Switzerland: nationally representative cross sectional study. Bmc Public Health. 2014; 14.

8. Meltzer H, Bebbington P, Dennis MS, Jenkins R, McManus S, Brugha TS. Feelings of loneliness among adults with mental disorder. Social Psychiatry and Psychiatric Epidemiology. 2013; 48(1): 5–13.

9. Liebke L, Bungert M, Thome J, Hauschild S, Gescher DM, Schmahl C, et al. Loneliness, Social Networks, and Social Functioning in Borderline Personality Disorder. Personality Disorders-Theory Research and Treatment. 2017; 8(4): 349–56.

10. Lim MH, Gleeson JFM, Alvarez-Jimenez M, Penn DL. Loneliness in psychosis: a systematic review. Social Psychiatry and Psychiatric Epidemiology. 2018; 53(3): 221–38.

11. Miret M, Caballero FF, Huerta-Ramirez R, Moneta MV, Olaya B, Chatterji S, et al. Factors associated with suicidal ideation and attempts in Spain for different age groups. Prevalence before and after the onset of the economic crisis. Journal of Affective Disorders. 2014; 163: 1–9.

12. VanderWeele TJ, Hawkley LC, Cacioppo JT. On the Reciprocal Association Between Loneliness and Subjective Well-being. Am J Epidemiol. 2012; 176(9): 777–84.

13. Gariepy G, Honkaniemi H, Quesnel-Vallee A. Social support and protection from depression: systematic review of current findings in Western countries. British Journal of Psychiatry. 2016; 209(4): 286–95.

14. Ma R. The role of subjective and objective social isolation as predictors of mental health recovery. In: Psychiatry. UCL, 2020.

15. Erzen E, Cikrikci O. The effect of loneliness on depression: A meta-analysis. International Journal of Social Psychiatry. 2018; 64(5): 427–35.

16. Wang J, Mann F, Lloyd-Evans B, Ma R, Johnson S. Associations between loneliness and perceived social support and outcomes of mental health problems: a systematic review. BMC Psychiatry. 2018; 18(1): 156.

17. Pluye PR, E.; Cargo M., Bartlet, G.; O’Cathain, A; Griffiths, F; Boardman, F.; Gagnon, M.P.; Rousseau, M.C.. Proposal: a mixed methods appraisal tool for systematic mixed studies reviews. 2011.

18. Wells GSB, O’Connell DO; Peterson J; Welch V, Losos M; Tugwell P. The Newcastle-Ottawa Scale (NOS) for assessing the quality of nonrandomised studies in meta-analyses. University of Ottawa. http://www.ohri.ca/programs/clinical_epidemiology/oxford.asp.

19. Popay JR, H.; Sowden, A.; Petticrew, M.; Arai, L.; Rodgers, M.; Britten, N.; Roen, K.; Duffy, S. Guidance on the conduct of narrative synthesis in systematic reviews: a product from the ESRC Methods Programme. ESRC, 2006.

20. Richardson TE, P.; Roberts, R. Relationship between loneliness and mental health in students. Journal of Public Mental Health. 2017; 16(2): 48–54.

21. Flensborg-Madsen T, Tolstrup J, Sorensen HJ, Mortensen EL. Social and psychological predictors of onset of anxiety disorders: results from a large prospective cohort study. Soc Psychiatry Psychiatr Epidemiol. 2012; 47(5): 711–21.

22. Vicente FE-S, H.; Cardoso, D.; da Silva, F.; Costa, M.; Martins, S.; Torres-Pea, Ines; Pascoal, V.; Rodrigues, F.; Pinto, A; Moitinho, S.; Guadalupe, S., Vicente, HT; Lemos, L. Longitudinal study of factors associated with the developement of depressive symptoms in institutionalized elderly. Jornal Brasileiro de Psiquiatria. 2014; 63(4).

23. Smalbrugge M, Jongenelis L, Pot AM, Eefsting JA, Ribbe MW, Beekman AT. Incidence and outcome of depressive symptoms in nursing home patients in the Netherlands. Am J Geriatr Psychiatry. 2006; 14(12): 1069–76.

24. Sjoberg L, Ostling S, Falk H, Sundh V, Waern M, Skoog I. Secular changes in the relation between social factors and depression: a study of two birth cohorts of Swedish septuagenarians followed for 5 years. J Affect Disord. 2013; 150(2): 245–52.

25. Green BH, Copeland JR, Dewey ME, Sharma V, Saunders PA, Davidson IA, et al. Risk factors for depression in elderly people: a prospective study. Acta Psychiatr Scand. 1992; 86(3): 213–7.

26. Prince MJ, Harwood RH, Thomas A, Mann AH. A prospective population-based cohort study of the effects of disablement and social milieu on the onset and maintenance of late-life depression. The Gospel Oak Project VII. Psychol Med. 1998; 28(2): 337–50.

27. Stessman J, Rottenberg Y, Shimshilashvili I, Ein-Mor E, Jacobs JM. Loneliness, health, and longevity. J Gerontol A Biol Sci Med Sci. 2014; 69(6): 744–50.

28. Beutel ME, Brahler E, Wiltink J, Kerahrodi JG, Burghardt J, Michal M, et al. New onset of depression in aging women and men: Contributions of social, psychological, behavioral, and somatic predictors in the community. Psychological Medicine. 2019; 49(7): 1148–55.

29. Conde-Sala JL, Garre-Olmo J, Calvo-Perxas L, Turro-Garriga O, Vilalta-Franch J. Course of depressive symptoms and associated factors in people aged 65+ in Europe: A two-year follow-up. Journal of Affective Disorders. 2019; 245: 440–50.

30. Cacioppo JT, Hughes ME, Waite LJ, Hawkley LC, Thisted RA. Loneliness as a specific risk factor for depressive symptoms: cross-sectional and longitudinal analyses. Psychol Aging. 2006; 21(1): 140– 51.

31. Cacioppo JT, Hawkley LC, Thisted RA. Perceived social isolation makes me sad: 5-year cross-lagged analyses of loneliness and depressive symptomatology in the Chicago Health, Aging, and Social Relations Study. Psychol Aging. 2010; 25(2): 453–63.

32. Theeke LA. Sociodemographic and health-related risks for loneliness and outcome difference by loneliness status in a sample of US older adults. In: Nursing. West Virginia University, 2007.

33. Luo Y, Hawkley LC, Waite LJ, Cacioppo JT. Loneliness, health, and mortality in old age: a national longitudinal study. Soc Sci Med. 2012; 74(6): 907–14.

34. Russell DW. UCLA Loneliness Scale (Version 3): reliability, validity, and factor structure. J Pers Assess. 1996; 66(1): 20–40.

35. Prince MJ, Harwood RH, Thomas A, Mann AH. A prospective population-based cohort study of the effects of disablement and social milieu on the onset and maintenance of late-life depression. The Gospel Oak Project VII. Psychological Medicine. 1998; 28(2): 337–50.

36. Green BH, Copeland JRM, Dewey ME, Sharma V, Saunders PA, Davidson IA, et al. Risk-Factors for Depression in Elderly People - a Prospective-Study. Acta Psychiatrica Scandinavica. 1992; 86(3): 213–7.

37. Vicente FE-S HC, D.; da Silva, F.; Costa, M.; Martins, S.; Torres-Pea, Ines; Pascoal, V.; Rodrigues, F.; Pinto, A; Moitinho, S.; Guadalupe, S., Vicente, HT; Lemos, L. Longitudinal study of factors associated with the developement of depressive symptoms in institutionalized elderly.. Jornal Brasileiro de Psiquiatria. 2014; 63(4).

38. Domenech-Abella J, Mundo J, Haro JM, Rubio-Valera M. Anxiety, depression, loneliness and social network in the elderly: Longitudinal associations from The Irish Longitudinal Study on Ageing (TILDA). Journal of Affective Disorders. 2019; 246: 82–8.

39. Richardson TE PR, R. Relationship between loneliness and mental health in students. Journal of Public Mental Health. 2017; 16(2): 48–54.

40. Nuyen J, Tuithof M, de Graaf R, van Dorsselaer S, Kleinjan M, ten Have M. The bidirectional relationship between loneliness and common mental disorders in adults: findings from a longitudinal population-based cohort study. Social Psychiatry and Psychiatric Epidemiology. 2020; 55(10): 1297– 310.

41. Stessman J, Rottenberg Y, Shimshilashvili I, Ein-Mor E, Jacobs JM. Loneliness, Health, and Longevity. J Gerontol a-Biol. 2014; 69(6): 744–50.

42. Luo Y, Hawkley LC, Waite LJ, Cacioppo JT. Loneliness, health, and mortality in old age: A national longitudinal study. Social Science & Medicine. 2012; 74(6): 907–14.

43. Cacioppo JT, Hughes ME, Waite LJ, Hawkley LC, Thisted RA. Loneliness as a specific risk factor for depressive symptoms: Cross-sectional and longitudinal analyses. Psychology and Aging. 2006; 21(1): 140–51.

44. Biaggi A, Conroy S, Pawlby S, Pariante CM. Identifying the women at risk of antenatal anxiety and depression: A systematic review. Journal of Affective Disorders. 2016; 191: 62–77.

45. Luoma I, Korhonen M, Puura K, Salmelin RK. Maternal loneliness: Concurrent and longitudinal associations with depressive symptoms and child adjustment. Psychology, Health & Medicine. 2019; 24(6): 667–79.

46. Goosby BJ, Bellatorre A, Walsemann KM, Cheadle JE. Adolescent Loneliness and Health in Early Adulthood. Sociol Inq. 2013; 83(4).

47. Flensborg-Madsen T, Tolstrup J, Sorensen HJ, Mortensen EL. Social and psychological predictors of onset of anxiety disorders: results from a large prospective cohort study. Social Psychiatry and Psychiatric Epidemiology. 2012; 47(5): 711–21.

48. Michalska da Rocha B, Rhodes S, Vasilopoulou E, Hutton P. Loneliness in Psychosis: A Meta-analytical Review. Schizophr Bull. 2018; 44(1): 114–25.

49. Alasmawi KM, F; Lewis, G; Mezey, G; White, S; Lloyd-Evans, B. To what extent does severity of loneliness vary among different mental health diagnostic groups: A cross-sectional study. International Journal of Mental Health Nursing. 2020; 29(5): 921–34.

50. Qualter P, Vanhalst J, Harris R, Van Roekel E, Lodder G, Bangee M, et al. Loneliness Across the Life Span. Perspectives on Psychological Science. 2015; 10(2): 250–64.

51. Balter LJT, Raymond JE, Aldred S, Drayson MT, van Zanten JJCSV, Higgs S, et al. Loneliness in healthy young adults predicts inflammatory responsiveness to a mild immune challenge in vivo. Brain Behav Immun. 2019; 82: 298–301.

52. Lai JCL, Leung MOY, Lee DYH, Lam YW, Berning K. Loneliness and Diurnal Salivary Cortisol in Emerging Adults. Int J Mol Sci. 2018; 19(7).

53. van Winkel M, Wichers M, Collip D, Jacobs N, Derom C, Thiery E, et al. Unraveling the Role of Loneliness in Depression: The Relationship Between Daily Life Experience and Behavior. Psychiatry. 2017; 80(2): 104–17.

54. Paque KB, H; Van Bogaert, P; Dilles, T. Living in a nursing home: a phenomenological study exploring residents’ loneliness and other feelings. Scand J Caring Sci 2018; 32(4): 1477–84.

55. Mann F, Bone JK, Lloyd-Evans B, Frerichs J, Pinfold V, Ma R, et al. A life less lonely: the state of the art in interventions to reduce loneliness in people with mental health problems. Soc Psychiatry Psychiatr Epidemiol. 2017; 52(6): 627–38.

56. Masi CM, Chen HY, Hawkley LC, Cacioppo JT. A meta-analysis of interventions to reduce loneliness. Pers Soc Psychol Rev. 2011; 15(3): 219–66.

57. de Jong Gierveld JvT, T.G.; Dykstra, P.A. Loneliness and Social Isolation. In: The Cambridge Handbook of Personal Relationships (ed AP Vangelisti, D.). Cambridge University Press, 2016.

58. Beller J, Wagner A. Loneliness and Health: The Moderating Effect of Cross-Cultural Individualism/Collectivism. J Aging Health. 2020; 32(10): 1516–27.

