## Supplementary Material 1 Search Terms for "Loneliness and the onset of new mental health problems in the general population: a systematic review"

**List of search terms**

Examples of MEDLINE search terms used in initial search for this review, adapted as required for other databases. Search conducted for articles on loneliness AND mental health problems AND onset-related terms.

### Loneliness and related terms

‘Loneliness’ OR ‘lonely’ OR ‘social support adj5 personal or perceived or quality’ (i.e. the phrase ‘social support’ with the subsequent terms within five words of it OR ‘confiding relationship*’)

### Mental health problem terms

Search terms included mental disorders [MeSH]. exp OR mental OR psychiatry* OR schizo* OR psychosis OR psychotic OR depress* OR mania* OR manic OR bipolar adj5 (disorder or disease or illness (OR anxiety disorders [MeSH].exp

### Onset related terms

Terms covering ‘onset’ of mental health problems included ‘onset OR ‘first-episode’ OR incidence [MeSH]
