## Supplementary Material 2 Characteristics Table Detail for "Loneliness and the onset of new mental health problems in the general population: a systematic review"

| **Author**  **Year**  **Country** | **Quality**  **rating^a^** | **Sample size and characteristics** | **Measures (predictor and outcome)** | **Length of follow-up** | **Covariates adjusted for** | **Results^b^**  **++, +, -** | **Statistical analysis and main results** | **Comments** |
| --- | --- | --- | --- | --- | --- | --- | --- | --- |
| **Depression – ‘pure onset’ studies** | | | | | | | | |
| Beutel (2018) Germany | *** | N=10,036  Age: 40-74 | Loneliness: single item "I am frequently alone / have few contacts" (recoded into 1 = no loneliness or distress; 2 = slight; 3 = moderate; and  4 = severe loneliness)  Depression: PHQ-9 (=/> 10) | Enrolled 2007  5-year follow-up | PHQ-9 at baseline; sociodemographic (sex, age, SES, partnership. psychological (type D, life events, social support, loneliness, GAD-2>3, social phobia, panic, history of AD), behavioural (active sports, obesity, smoking, alcohol abuse), somatic (CVD, COPD, cancer, diabetes) | ++ | Logistic regression analysis  Baseline loneliness and social support were both associated with onset of depression at 5 years:  Model 1 (excluding PHQ-9 at baseline model):  loneliness aOR 2.012 (1.479, 2.709),  social support aOR 0.926 (0.900, 0.954)  model 2 (including PHQ-9 at baseline): loneliness aOR 1.551 (1.135, 2.099)  social support aOR 0.954 (0.927, 0.984) | Excluded subjects with medical history of depression, intake of antidepressant medication, and increased depression scores (PHQ>10) at baseline |
| Conde-Sala (2019) Spain | *** | N = 31,491  Older adults over 65 | Loneliness: Hughes et al., 2004 3-item loneliness scale (>3/9 = loneliness)   Depression: 12-item EURO-D (≥4/12 = clinically relevant depressive symptoms) | Baseline 2013  2-year follow-up | Age, gender, schooling, financial difficulties, self-rated health, chronic diseases, ADL impairment, cognition | ++ | Multivariate binary logistic regression analyses  Loneliness at baseline significantly associated with new incidence of clinically depressive symptoms at follow-up  Loneliness associated with depression  Incidence (vs no-depression) aOR 1.63 (1.62, 1.64) Persistence (vs non-depression) aOR 3.10 (3.09, 3.11) Remission (vs persistence) OR 1.39 (1.38, 1.39) |  |
| Green^36^  (1992)  UK | * | N=1070  Older people over 65 | Loneliness: single item (do you feel lonely? 0=disagree 1=agree 2=strongly agree)  Depression: AGECAT diagnosis based on Geriatric Mental State data) | Enrolled 1982-3  3-year follow-up | Log-linear modelling tested independence of risk factors including age, ethnicity, alcohol use, bereavement, contact with friends or relatives, past psychiatric history | + | ‘New depression’ group was compared with those with no depression at follow-up.  New depression group significantly more lonely at baseline (chi-squared 16.98, p<0.0005)  Odds of loneliness at baseline in ‘new depression’ group significantly different (OR 1.82) | Living alone was NOT useful predictor of depression  No confidence interval available for OR (communication from author) |
| Prince^37^  (1998)  UK | *** | N=538  Older people over 65 | Loneliness: single item ‘often feeling lonely’  Depression (SHORT-CARE scale) | Enrolled 1993  1-year follow-up | Age, sex, five social support domains were not significant predictors of depression | ++ | Risk of new onset depression higher in people who often felt lonely RR 3.6 (2.0-6.4) | Other domains of social support were NOT significant predictors of depression. Also no significant association with age or sex. |
| Sjoberg^35^ (2013)  Sweden | *** | N=245 (1901 birth cohort)  N=310 (1930 birth cohort)  Older people (recruited aged 70), general population | Loneliness: single item (seldom/never vs sometimes/often)  Depression: DSM-IV diagnosis | 2 birth cohorts    5 years follow-up each | Sex, marital status | ++ | Baseline loneliness was associated with onset of depression at 5 years  1901 births:  aOR 3.81 (1.10-13.20)  1930 births:  aOR 2.83 (1.23-6.39) | The more objective variable ‘contact with others’ (quantity) was only associated with depression onset in older cohort |
| Stessman^38^  (2014)  Israel | ** | N=340 (1990 recruited cohort)  N=705 (1998 recruited cohort)  Two cohorts:   1) Age 70-78 2) Age 78-85 | Loneliness: single item (never lonely vs rarely/often/very often lonely)  Depression: Brief Symptoms Inventory | 2 cohorts  7 years follow-up each | Sex, marital status, education, self-rated health, physical activity, chronic pain, hypertension, ischaemic heart disease, diabetes | -  (+) | ‘Never lonely’ vs ‘any loneliness’ variable was NOT associated with new depression:  Age 70-78 aOR 0.61 (0.137-2.68)  Age 78-85 aOR 1.61 (0.8-3.25)  Categorising as ‘never/rarely lonely’ vs ‘often or very often’ at age 78 predicts new depression at 85. aOR 2.42 (1.18-4.9) | Data also analysed separately for men and women: no change to overall results. Loneliness not associated with mortality or other physical health outcomes either |
| Smalbrugge  (2006)^34^  Netherlands | *** | N=218  Older people over 55 (48% under 80) | Loneliness: De Jong Gerveld loneliness scale (>3/11 = ‘highly lonely’)  Depression: Geriatric Depression Scale | Enrolled 1999-2001  6 months follow-up | Age, gender, urbanisation (area), depressive symptoms, pain, functional limitations, stroke, perceived inadequacy of care | - | Loneliness NOT associated with onset of depression  Unadjusted OR 0.07 (0.01-0.57) | Very small number of people with new depression (n=10) meant adjusted OR could not be calculated  (communication from author) |
| **Depression onset and outcome** | | | | | | | | |
| Cacioppo^39^  (2006)  USA | *** | Older people, aged 50-68  N=212  *CHASRS* cohort* | Loneliness: UCLA loneliness scale  Depression: CES-D (minus loneliness item) | Baseline 2002, 3-year follow-up | Baseline depression, stress, social support, hostility, year of study, sex, ethnicity, age, marital status, education, income | ++ | Latent growth curve modelling  Baseline loneliness (year 1) predicts subsequent depression (coefficient 1.40, SE 0.55, p<0.05) | Depression predicts loneliness as well (1.56, SE 0.7, p<0.05)  Loneliness appeared fairly stable over the three years overall  *CHASRS cohort* |
| Cacioppo^40^  (2010)  USA | *** | Older people, aged 50-68  N=229  *CHASRS* cohort* | Loneliness: UCLA loneliness scale  Depression: CES-D (minus loneliness item) | Annual follow-up 2002-6 | Age, gender, marital status, race/ethnicity, antidepressant use, diagnosis, physical functioning | ++ | Significant 1-year cross-lagged effect of loneliness on depressive symptoms B=0.18 (0.09-0.30) across 5 years  Loneliness stable over time | Effect of loneliness was independent of demographic, health, and medication  Mix of ethnicities sampled including non-Hispanic White, Black and non-Black Latino Americans) |
| Goosby^48^  (2013)  USA | ** | N=10564  Nationally representative sample of high school students (18+) | Loneliness (items combined from CES-D, not validated)  Depression: CES-D | Wave 1  1994  Wave 2  1996 | Ethnicity, age, parent education, parent income, parent marital status, respondent nativity status, health insurance access, parent self-rated health, parent-reported respondent health at WAVW 1, region of residence, binge drinking frequents, regular smoking | ++ | Loneliness associated with new onset of depression  aOR 1.41 (1.35-1.50)  unadjusted OR 1.45, p<0.001  Parental support across baseline and follow-up moderates the depression between loneliness and depression aOR 1.25 (1.14-1.42) | Significant interaction between loneliness and parental support |
| Lim^49^  (2011)  Singapore | * | N=2799  Older people, mean age 66 (55+) | Loneliness: single item dichotomised not at all lonely vs fairly lonely/very lonely  Depression: Geriatric Depression Scale Score (GDS) | 2- year follow-up | Age, gender, race, marital status, education, social contact frequency, no. of medical problems, no. of social/productive/fitness/health activities, functional disabilities, cognitive status, baseline depression and QoL | ++ | Loneliness significant predictor of higher depression scores after 2 years  aOR 1.39, B=0.33, SE 0.36, p=0.03 | Loneliness was greater contributor to model of depressive symptoms (F13.91, p<0.001 than ‘living alone’ (F1.84, P = 0.18) |
| Ye Luo^42^  (2012)  USA | *** | N=2101  Older people, mean age 67  *Subset of HRS cohort* | Loneliness: 3-item UCLA loneliness scale  Depression: CES-D minus ‘I feel lonely’ and sleep items | 3 waves: 2002, 2004, 2006  2-year follow-ups | Marital status, relatives/friends nearby, sleep, exercise, smoking, age, gender, ethnicity, education, household income and assets | ++ | Significant 2-year cross-lagged effect of loneliness on depression  B=0.132, p<0.001 | Reciprocal effect of depression on loneliness over two years:  B=0.113, P<0.001 |
| Theeke^41^  (2007)  (thesis) | **** | N= 13 812  Older people (50+)  *HRS cohort* | Loneliness: single item from within CES-D (‘feeling lonely for most of past week). ‘Never lonely’ vs ‘briefly lonely’ vs ’chronically lonely’  Depression (CES-D 7 items) | Waves 2002-4  2 years | Independent analysis of covariance tests to control for marital status, health, education, functional statues, chronic illness, age, income, no. of people in household | + | Never lonely vs chronically lonely: mean difference in depression score 1.55 (error 0.03 p<0.005)  Briefly lonely vs chronically lonely mean difference in depression score 0.72 (error 0.04) p<0.005 | Analysis of covariance showed results for loneliness on depression remained significant |
| Vicente^33^  (2014)  Portugal | ** | N=83  Older people (institutionalised; average age 79.5) | Loneliness: UCLA loneliness scale  Depression: GDS | 2011-2013  2-year follow-up |  | - | Those whose depression scores worsened over time (including people who had no depression at baseline and people who had depression) had higher loneliness scores at baseline, but did not reach statistical significance | Of those whose loneliness scores worsened over time, a significantly greater proportion had worsening depression scores (compared with people whose depression scores improved or they remained depression-free). |
| Luoma^47^  (2015)  Finland | *** | N=329  Mothers (recruited first trimester) | Loneliness: single item (always/often/sometimes vs rarely/never)  Postnatal depression (Edinburgh Postnatal Depression Score) | Baseline 1989-90  16 to 17-year follow-up | Mother’s age, past or current mental health problems, EPDS score, ever smoked, relationship changed, ‘not very good’ pregnancy, difficulties during pregnancy, negative expectations | ++ | Group-based modelling identified a four cluster model was best for predicting depression trajectories (‘high stable’/’intermittent’/low stable’ and ‘very low’)  Feeling lonely associated with high stable depression symptoms. aOR 2.1 (1.0-4.2) p 0.041 | Feeling lonely was not associated with ‘intermittent’ trajectory |
| Richardson  (2017)^43^  UK | ** | N=454 (2 cohorts combined)  University students  Mean age 19.9 | Loneliness: 3-item UCLA loneliness scale  CES-D | 2012-2014; 2 cohorts followed up over 12-14 months  First cohort (baseline Feb-June 2012): FU at 3/12, 6/12 and 12/12  Second cohort (baseline Oct/Dec 2012): FU at 3/12 and 6/12, 12/12 | Age, gender, ethnicity, baseline scores | + | Baseline loneliness correlated with depression (time 2 r=0.51, time 3 r=0.48, time 4 r=0.42) p<0.001  After accounting for demographics and baseline scores, loneliness predicted depression only at T4 (beta=0.14, p<0.05) | No evidence that presence of mental health problems predicted increased loneliness over time  Sample nearly 80% female |
| Domenech-Abella (2019)  Ireland | **** | N = 5066  Community-dwelling adults aged 50 years and older in Ireland  Mean age at baseline: 63.3 | Loneliness: 5-item UCLA loneliness scale (hardly ever or never (1) to often (2)), range from 0-10  Composite International Diagnostic Interview-Short Form (CIDI-SF) to assess MDD in past 12 months | 2009-2011 until 2014-2015; 3 waves  5-6 year-follow-up | Sociodemographic characteristics (age, sex, education, financial circumstances, widowhood, employment status, place of residence), heart diseases, somatic diseases, affective disorders, social network index (SNI) | ++ | After adjusting for covariates, loneliness at wave 2 predicted depression at wave 3: aOR 1.22 (1.15-1.30) |  |
| **Anxiety** | | | | | | | | |
| Flensborg-Madsen^44^  (2012)  Denmark | *** | N=4497  Adults (mean age 44.9) | Loneliness: single item (no, in doubt, yes)  Anxiety disorder presence (national registers ICD8: 300, 300.2, 300.3, ICD-10: F40-43) | Enrolled 1993  13-year-follow-up | Age, yearly income, number of diseases at times of investigation | ++ | Multiple Cox regression analysis  Women: ‘Yes’ vs ‘no’ lonely and being hospitalized with anxiety: HR 2.01 (1.31-3.06)  ‘in doubt’ vs ‘no’ HR 1.14 (0.64-2.01)  Men: ‘Yes’ vs ‘no’ lonely and later being hospitalized with anxiety: HR 2.34 (1.34-4.09)  ‘in doubt’ vs ‘no’ HR 2.03 (1.19-2.63) |  |
| Richardson  (2017)^43^  UK | ** | N=454 (2 cohorts combined)  University students  Mean age 19.9 | Loneliness: 3-item UCLA loneliness scale  CES-D | 2012-2014; 2 cohorts followed up over 12-14 months  First cohort (baseline Feb-June 2012): FU at 3/12, 6/12 and 12/12  Second cohort (baseline Oct/Dec 2012): FU at 3/12 and 6/12, 12/12 | Age, gender, ethnicity, baseline scores | + | Baseline loneliness correlated with anxiety time 2 (r=0.41, T3 r=0.40, T4 r=0.34) p<0.001  After accounting for demographics, loneliness predicted anxiety only at T3 (beta 0.15, p<0.01) | Study looked at depression , anxiety, and ‘core mental health’ (below) |
| Domenech-Abella (2019)  Ireland | **** | N = 5066  Community-dwelling adults aged 50 years and older in Ireland  Mean age at baseline: 63.3 | Loneliness: 5-item UCLA loneliness scale (hardly ever or never (1) to often (2)), range from 0-10  Composite International Diagnostic Interview-Short Form (CIDI-SF) to assess GAD lasting six months or longer | 2009-2011 until 2014-2015; 3 waves  5-6 year-follow-up | Sociodemographic characteristics (age, sex, education, financial circumstances, widowhood, employment status, place of residence), heart diseases, somatic diseases, affective disorders, social network index (SNI) | ++ | After adjusting for covariates, loneliness at wave 2 predicted anxiety at wave 3: aOR 1.60 (1.10-2.34) |  |
| ‘**Core mental health’** |  |  |  |  |  |  |  |  |
| Richardson  (2017)^43^  UK | ** | N=454 (2 cohorts combined)  University students  Mean age 19.9 | Loneliness: 3-item UCLA loneliness scale  Core mental health:’ CORE-GP’ | 2012-2014; 2 cohorts followed up over 12-14 months  First cohort (baseline Feb-June 2012): FU at 3/12, 6/12 and 12/12  Second cohort (baseline Oct/Dec 2012): FU at 3/12 and 6/12, 12/12 | Age, gender, ethnicity, baseline scores | + | After adjusting for demographics and baseline scores, loneliness predicted core mental health at T2 (beta 0.11, p<0.05). |  |
| Nuyen (2019)  Netherlands | *** | N=4007  General population aged 18-64  Mean age: 44.3 years | Loneliness: De Jong Gerveld loneliness scale  Common mental disorders: Composite International Diagnostic Interview (CIDI) version 3.0 | Baseline: 2013-2015  3-year follow-up | gender, age, education, living situation, job status, household income, recent negative life event, perceived social support | ++ | After adjusting for covariates, loneliness at wave 2 predicted the onset of severe 12-month CMD at wave 3;  aRRR 3.28 (1-54-7.02)  After adjusting for covariates, loneliness at wave 2 did not predict onset of a mild-moderate 12-month CMD;  aRRR 0.94 (0.50-1.17) |  |

a Detailed quality ratings in Appendix 2

b ++ p<0.05, adjusted ; + p<0.05 unadjusted, - non-significant

c both drawn from same larger Chicago Health, Aging and Social Relations study, but different statistical approaches, and follow-up, d Health and Retirement Study

**Supplementary material Table 1 Characteristics and main findings from included studies across depression, anxiety and ‘core mental health’**
