## Supplementary Material 3 Quality Ratings for "Loneliness and the onset of new mental health problems in the general population: a systematic review"

Quality assessment

**Supplementary Material 2** Individual quality ratings: studies focusing on onset and outcome of mental disorders using Mixed Methods Appraisal Tool (MMAT)

| 1st author, publication year | **Screening questions** | | Selection bias | Measurements | Groups comparable | Outcome data/response rate/follow-up rate | Overall score |
| --- | --- | --- | --- | --- | --- | --- | --- |
|  | **Research questions** | **Collected data** |  |  |  |  |  |
| **Depression** | | | | | | | |
| Beutel, 2018 | Yes | Yes | Yes (stratified random sampling for gender, residence, age) | No (1 item for loneliness) | Yes | Yes (98.1% baseline response rate and 82.80% follow-up rate (of which 97.7% filled out the PHQ-9)) | *** |
| Cacioppo, 2006 | Yes | Yes | Yes | Yes | Yes | NO (45% baseline response rate) | *** |
| Cacioppo, 2010 | Yes | Yes | Yes | Yes | Yes | No (response rate < 60%) | *** |
| Conde-Sala, 2018 | Yes | Yes | Yes | Yes | Yes | No (no information on baseline response rate and 26.3% follow-up attrition rate) | *** |
| Goosby, 2013 | Yes | Yes | Yes | No (not a validated tool) | Yes | Can’t tell (no response rate and follow-up rate) | ** |
| Green, 1992 | Yes | Yes | Yes | No (1 item for loneliness) | Yes | Can’t tell (no % of complete data and follow-up rate, lack of confidence intervals for risk estimate) | ** |
| Harris 2006 | Yes | Yes | No | No (single item loneliness, but GDS-15 depression) | Yes | No - baseline response rate 75%, follow-up response rate 94% | * |
| Lim, 2011 | Yes | Yes | Can’t tell (not clear enough how they selected their sample) | No (1 item for loneliness) | Yes | Can’t tell (no % of complete data) | * |
| Luoma, 2015 | Yes | Yes | Yes | Yes | Yes | No – baseline response rate 90%, but large dropout at FUs | *** |
| Prince, 1998 | Yes | Yes | Yes | Yes | Yes | No (completeness of outcome data) | *** |
| Sjoberg, 2013 | Yes | Yes | Yes | No (1 item for loneliness) | Yes | Yes | *** |
| Smallbrugge, 2006 | Yes | Yes | Yes | Yes | Yes | No | *** |
| Stessman, 2014 | Yes | Yes | Yes | No (1 item for loneliness) | Yes | Can’t tell (no response rate and follow-up rate) | ** |
| Theeke,  2007 | Yes | Yes | Yes | No (single item loneliness, but CES-D depression) | Yes | Yes (baseline response rate 87%) | **** |
| Vincente 2014 | Yes | Yes | No (several people not included/assessed down to practical issues like location and lack of resources to visit etc) | Yes | Yes | Original response rate not clear, completeness of data not clear  No | ** |
| Ye Luo, 2012 | Yes | Yes | Yes | Yes | Yes | Can’t tell (no % of complete outcome data) | *** |
| **Anxiety disorders** | | | | | | | |
| Flensborg-Madsen, 2012 | Yes | Yes | Yes | No (1 item for loneliness and for quality of social network) | Yes | Yes | *** |
| **Mixed mental disorders (depression, anxiety, core mental health)** | | | | | | | |
| Domènech-Abella 2019 | Yes | Yes | Yes | Yes | Yes | No  62% baseline response;  86% wave 2 response rate;  85% wave 3 response rate | *** |
| Nuyen 2019 | Yes | Yes | Yes | Yes | Yes | *No*  Response rate from T0 to T3 61.6% but response rate from T2 to T3 (data on these time points analysed): 86.8% | *** |
| Richardson, 2017  UK | Yes | Yes | No – limited info on recruitment methods | Yes | Yes | Response rate not given at baseline. Just under 50% lost at follow-up Time 4. Data completion adequate, and mode substituted for missing values. | ** |

Newcastle-Ottawa Quality Assessment Scale

| 1st author, publication year | Representativeness of the exposed cohort | Demonstration that outcome of interest was not present at start of study |
| --- | --- | --- |
|  | a) truly representative of the average _______ (describe) in the community *  b) somewhat representative of the average _______ in the community *  c) selected group of users e.g. nurses, volunteers  d) no description of the derivation of the cohort | a) yes *  b) no |
| **Depression** | | |
| Beutel, 2018 | b) somewhat representative of the average person between the age of 35 and 74 in the city of Mainz and the district of Mainz-Bingen* | a) yes * |
| Cacioppo, 2006 | b) somewhat representative of the average level of loneliness and perceived social support in a sample of European American, African American and Latino American aged 50-67 in Cook County, Illinois * | a) yes* |
| Cacioppo, 2010 | b) feeling of loneliness * | b) no |
| Conde-Sala, 2018 | a) truly representative of the average older person over 65 from 14 European countries* | a) yes * |
| Goosby, 2013 | c) high school students | b) no |
| Green , 1992 | a) feeling of loneliness in people aged 65 years and above * | a) yes * |
| Harris 2006 | b) somewhat representative of the average older person over 65 in Southeast London | Yes* - included follow-up group that had no depression at baseline |
| Lim, 2011 | d) | b) no |
| Luoma, 2015 | b) somewhat representative of the average adult expectant mother in the community* | No – those with few symptoms and NO symptoms grouped together |
| Prince, 1998 | a) truly representative of the average elderly person with disablement and loneliness in the community* | Yes* |
| Sjoberg, 2013 | a) feeling of loneliness * | a) yes * |
| Smallbrugge, 2006 | b) somewhat representative of depression in the average elderly nursing home resident * | a) yes (GDS <10 at baseline)* |
| Stessman, 2014 | a) feeling of loneliness * | a) yes * |
| Theeke 2007 | b) somewhat representative of the average older person over 50 in the US* | no |
| Vicente, 2014 | c) institutionalized elderly, without organic brain or cognitive impairment | *yes – included one follow-up group that had no depression at baseline and remained that way (used GDS cut-off) but study overall included onset and outcomes |
| Ye Luo, 2012 | a) feeling of loneliness * | b) no |
| **Anxiety disorders** | | |
| Flensborg-Madsen, 2012 | a) feeling of loneliness and quality of social network * | a) yes * |
| **Mixed mental disorders** | | |
| Richardson, 2017 | c) selected group of students in the UK | no – though baseline scores were adjusted for in regression analysis |
| Nuyen 2019 | a) Truly representative of the average 18-64-year old person in the Netherlands (however, younger subjects somewhat underrepresented) | yes* - included follow-up group that had no 12-month CMD at baseline |
| Domènech-Abella 2019 | a) Truly representative of the average older person over 50 in Ireland | no - though baseline scores were adjusted for in regression analysis |
