## Supplementary Material 4 Forest Plot Continuous for "Loneliness and the onset of new mental health problems in the general population: a systematic review"

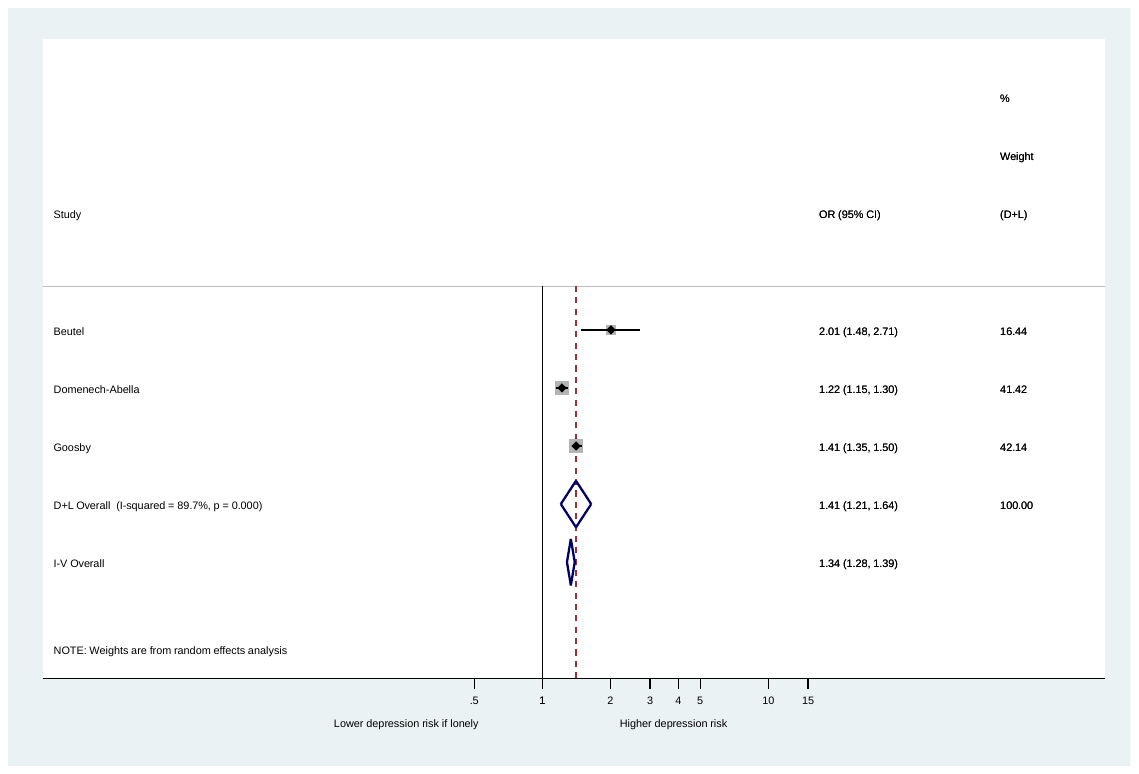

Supplementary Figure 4

Forest Plot to show association between baseline loneliness and onset of depression (continuous loneliness measures).
