## Supplementary Material 5 Depression and Loneliness Rates for "Loneliness and the onset of new mental health problems in the general population: a systematic review"

**Supplementary Material 5** Information on rates of depression and loneliness as provided by studies

| First author | Loneliness | Depression |
| --- | --- | --- |
| Sjoberg (2013) | Women:  1901 cohort, 23.4% loneliness at baseline  1930 cohort, 32.2% loneliness at baseline  Men:  1901 cohort, 11.9% loneliness at baseline  1930 cohort, 17.5% loneliness at baseline | Women:  1901 cohort 7.6% rate of new depression  1930 cohort 18.2% rate of new depression  Women:  1901 cohort 7.9% rate of new depression  1930 cohort 9.7% rate of new depression |
| Stessman (2014) | Baseline loneliness age 70: 28%, age 78: 24%, age 85: 24% | New depression age 70-78: 8.1%  New depression age 78-85:  22.3% |
| Smallbrugge (2006) | Loneliness at baseline: 64.8% | New depression: 4.7% |
| Green (1992) | Loneliness at baseline: not given | New depression: 4.1% |
| Prince (1998) | Loneliness at baseline not given, 23% of people with new depression were lonely | New depression: 12% |
| Goosby (2013) | Loneliness at baseline given as mean score on scale made by authors using composite of items on various scales featuring ‘loneliness-related’ items: -0.01 (range -1.03 to 5.6), SE 0.83 | Overall depression at follow-up: 10% |
| Lim (2011) | Loneliness at baseline: 11.9% | Depression rate overall not given |
| Cacioppo (2010) | Rate not given | Rate not given |
| Cacioppo (2006) | Loneliness mean score at baseline (UCLA loneliness scale) 36.0, SD 9.9 | Mean depression score only given at baseline (CES-D minus ‘lonely’ item) 9.6 (SD 8.5) |
| Ye Luo (2012) | Loneliness at baseline: three two-year cohorts (score range 3-9). Mean scores were 2002: 3.84, 2004: 3.8, 2006: 4.29 | Depression scores (modified CES-D for use over telephone with elderly, range 0-6) 2202: 0.95, 2004: 0.89, 2006: 0.90 |
| Theeke (2007) | Loneliness 16.9%, 8.8% ‘chronically lonely’ | Depression rates not given |
| Vicente (2014) | Mean UCLA loneliness score in people who were not depressed at baseline: 32.6 +/-10.56; score in people that were depressed at baseline (GDS>10) : 38.2+/-10.41 | New depression: 10.8% |
| Luoma (2015) | Percentage ‘always, often or sometimes lonely’) in different trajectory groups: very low depression throughout: 12%, low-stable 39%, high-stable 58%, intermittent 30% | Different trajectories of depression: very low depression score throughout: very low-stable (i.e. started with low score and remained low at follow-up) 53%, high-stable 27%, intermittent 3% |
| Richardson (2017) | Loneliness rate not given | Depression/anxiety rate not given |
| Flensborg-Madsen (2012) | Baseline loneliness rate not given | Registered with anxiety disorder by follow-up 5.3% |
| Beutel (2019) | Loneliness at baseline: 6.6% in total,  6.1% without depression,  18.7% with depression | New onset depression: 4.4% |
| Conde-Sala (2018) | Loneliness at baseline (wave 5): 43.7% Loneliness at follow-up (wave 6): 47.5% | Overall depression at baseline (wave 5): 29.8%  Overall depression at follow-up (wave 6): 31.5% |
| Domenech-Abella (2019) | At T1 13.6% of the general population had a score of 5-10 on the loneliness scale, and out of these 9.10% exhibited worse, 26.50% consistent, and 64.40% improved symptoms in wave 3  At T1 86.4% of the general population had a score of 0-4 on the loneliness scale, and out of these exhibited 22.6% worse, 53.70% consistent, and 23.70% improved symptoms in wave 3 | 5.9% of the general population had MDD at T1, out of these 77.2% showed no change and 22.80% showed an improvement in symptoms in wave 3; 3.9% of the 94.1% without MDD at T1 showed worsening of symptoms in wave 3  2.8% of general population had GAD at T1, out of these 74.9% showed no change and 25.10% showed improved in symptoms in wave 3; 2.1% of the 97.2% without MDD at T1 showed worsening in wave 3 |
| Harris 2006 | Loneliness at baseline: 12% sometimes and 22.7% often/always | 8.4% rate of new depression |
| Nuyen 2019 | Loneliness at baseline: 38% overall and 16.8% of individuals without 12-month CMD at baseline | 5.8% rate of new mild-moderate CMD  2.8% rate of severe CMD  17.8% rate of new mild-moderate CMD and 45.6% rate of new severe CMD among individuals without CMD at baseline |
